# Red Teaming Large Language Models in Medicine: Real-World Insights on Model Behavior

**DOI:** 10.1101/2024.04.05.24305411

**Authors:** Crystal T. Chang, Hodan Farah, Haiwen Gui, Shawheen Justin Rezaei, Charbel Bou-Khalil, Ye-Jean Park, Akshay Swaminathan, Jesutofunmi A. Omiye, Akaash Kolluri, Akash Chaurasia, Alejandro Lozano, Alice Heiman, Allison Sihan Jia, Amit Kaushal, Angela Jia, Angelica Iacovelli, Archer Yang, Arghavan Salles, Arpita Singhal, Balasubramanian Narasimhan, Benjamin Belai, Benjamin H. Jacobson, Binglan Li, Celeste H. Poe, Chandan Sanghera, Chenming Zheng, Conor Messer, Damien Varid Kettud, Deven Pandya, Dhamanpreet Kaur, Diana Hla, Diba Dindoust, Dominik Moehrle, Duncan Ross, Ellaine Chou, Eric Lin, Fateme Nateghi Haredasht, Ge Cheng, Irena Gao, Jacob Chang, Jake Silberg, Jason A. Fries, Jiapeng Xu, Joe Jamison, John S. Tamaresis, Jonathan H Chen, Joshua Lazaro, Juan M. Banda, Julie J. Lee, Karen Ebert Matthys, Kirsten R. Steffner, Lu Tian, Luca Pegolotti, Malathi Srinivasan, Maniragav Manimaran, Matthew Schwede, Minghe Zhang, Minh Nguyen, Mohsen Fathzadeh, Qian Zhao, Rika Bajra, Rohit Khurana, Ruhana Azam, Rush Bartlett, Sang T. Truong, Scott L. Fleming, Shriti Raj, Solveig Behr, Sonia Onyeka, Sri Muppidi, Tarek Bandali, Tiffany Y. Eulalio, Wenyuan Chen, Xuanyu Zhou, Yanan Ding, Ying Cui, Yuqi Tan, Yutong Liu, Nigam H. Shah, Roxana Daneshjou

**Affiliations:** Department of Dermatology, Stanford University, Stanford, USA; Clinical Excellence Research Center, School of Medicine, Stanford University, Palo Alto, California; School of Medicine, Stanford University, Stanford, USA; Temerty Faculty of Medicine, Toronto, Canada; Department of Biomedical Data Science, Stanford University, Stanford, USA; Department of Computer Science, Stanford University, Stanford, USA; Department of Cardiothoracic Surgery, Stanford University, Stanford, USA; Stanford University, Stanford, USA; Department of Bioengineering, Stanford University, Stanford, USA; Department of Pediatrics, Stanford University, Stanford, USA; Department of Mathematics and Statistics, McGill University, Montreal, Canada; Department of Psychiatry, Stanford University, Stanford, USA; Mayo Clinic Alix School of Medicine, Rochester, USA; Department of Statistics, Stanford University, Stanford, USA; Veterans Affairs Medical Center, Palo Alto, USA; Center for Biomedical Informatics Research, Stanford University, Stanford, USA; Department of Anesthesiology, Stanford University, Stanford, USA; Department of Medicine, Stanford University, Stanford, USA; Stanford BioDesign, Stanford University, Stanford, USA; Department of Education and Psychology, Freie Universität Berlin, Berlin, Germany; Department of Epidemiology and Population Health, Stanford University, Stanford, USA; Technology and Digital Solutions, Stanford Health Care, Palo Alto, USA; Graduate School of Business, Stanford University, Stanford, USA; Department of Pathology, Stanford University, Stanford, USA

**Keywords:** Large language models, red teaming, bias in artificial intelligence, safety concerns in artificial intelligence, applications of artificial intelligence in medicine

## Abstract

0.

**Background:** The integration of large language models (LLMs) in healthcare offers immense opportunity to streamline healthcare tasks, but also carries risks such as response accuracy and bias perpetration. To address this, we conducted a red-teaming exercise to assess LLMs in healthcare and developed a dataset of clinically relevant scenarios for future teams to use.

**Methods:** We convened 80 multi-disciplinary experts to evaluate the performance of popular LLMs across multiple medical scenarios. Teams composed of clinicians, medical and engineering students, and technical professionals stress-tested LLMs with real world clinical use cases. Teams were given a framework comprising four categories to analyze for inappropriate responses: Safety, Privacy, Hallucinations, and Bias. Prompts were tested on GPT-3.5, GPT-4.0, and GPT-4.0 with the Internet. Six medically trained reviewers subsequently reanalyzed the prompt-response pairs, with dual reviewers for each prompt and a third to resolve discrepancies. This process allowed for the accurate identification and categorization of inappropriate or inaccurate content within the responses.

**Results:** There were a total of 382 unique prompts, with 1146 total responses across three iterations of ChatGPT (GPT-3.5, GPT-4.0, GPT-4.0 with Internet). 19.8% of the responses were labeled as inappropriate, with GPT-3.5 accounting for the highest percentage at 25.7% while GPT-4.0 and GPT-4.0 with internet performing comparably at 16.2% and 17.5% respectively. Interestingly, 11.8% of responses were deemed appropriate with GPT-3.5 but inappropriate in updated models, highlighting the ongoing need to evaluate evolving LLMs.

**Conclusion:** The red-teaming exercise underscored the benefits of interdisciplinary efforts, as this collaborative model fosters a deeper understanding of the potential limitations of LLMs in healthcare and sets a precedent for future red teaming events in the field. Additionally, we present all prompts and outputs as a benchmark for future LLM model evaluations.

**1-2 Sentence Description:** As a proof-of-concept, we convened an interactive “red teaming” workshop in which medical and technical professionals stress-tested popular large language models (LLMs) through publicly available user interfaces on clinically relevant scenarios. Results demonstrate a significant proportion of inappropriate responses across GPT-3.5, GPT-4.0, and GPT-4.0 with Internet (25.7%, 16.2%, and 17.5%, respectively) and illustrate the valuable role that non-technical clinicians can play in evaluating models.

## 1. Introduction

Large language models (LLMs) are a class of generative AI models capable of processing and generating human-like text at a large scale^1^. However, LLMs are susceptible to inaccuracies and biases in their training data. The objective of an LLM is to iteratively predict the next most likely word or word part. Because it does not necessarily reason through tasks, an LLM can produce “hallucinations,” or seemingly plausible utterances not grounded in reality. Additionally, popular models such as ChatGPT, GPT-4, Google Bard and Claude by Anthropic can all perpetuate racist tropes and debunked medical theories, potentially worsening health disparities^2^.

Despite these limitations, due to their vast promise, LLMs and other generative AI models are already being tested in the real-world clinical setting through high-profile partnerships first announced in the fall of 2023, such as the collaborations between leading electronic health record (EHR) vendors Epic and Oracle with Microsoft^3^ and Nuance^4^, respectively. Large technology companies like Microsoft and Google have also partnered with early adopter health systems, such as Mayo Clinic, Stanford, and NYU^5^. Providers are able to beta-test functions such as medical text summarization for automatic medical documentation generation, medical billing code suggestion, AI-drafted responses to patient messages, and more^1^.

While this represents a significant integration of potentially transformative technology, these announcements came less than a year after ChatGPT was released to the public in November 2022^6^, kick-starting a generative AI frenzy. Given the potential impact of generative AI on patient outcomes and public health, it is imperative that medicine, academia, government, and industry work together to address the challenges these models pose. To that end, in October 2023, President Biden issued the landmark ‘Executive Order on the Safe, Secure, and Trustworthy Development and Use of Artificial Intelligence’.^7^ The order, aimed at regulating and promoting the ethical development of AI systems, mandates that major AI developers must share all safety results, including the outcomes of red teaming tests, with the U.S. government before the systems are made publicly available.

Originally a cybersecurity term, red-teaming is the process of taking on the lens of an adversary (the ‘red team,’ as opposed to the defensive ‘blue team’) in order to expose system/model vulnerabilities and unintended or undesirable outcomes. These outcomes may include incorrect information due to model hallucination, discriminatory or harmful information or rhetoric, and other risks or potential misuses of the system. Red teaming can be done by software experts within the same firm, by rival firms, or by non-technical laypeople, such as when reddit users “jailbreak” LLM chatbots through prompts (input provided to models that then leads to a generated response) that bypass the models’ alignment^8^. Red teaming is critical to identifying flaws that can then be addressed and fixed using trustworthy AI, which are methods designed to test and strengthen the reliability of AI systems.

Though red teaming is a recognized and now federally mandated practice in the field of computer science, it is not well-known in healthcare. Yet clinician knowledge of prompts that are likely to be used for LLMs in the healthcare setting and evaluation of LLM response appropriateness are imperative towards robust evaluation of these models pre- and post-deployment. Furthermore, to minimize conflict of interest, it is important that people working in medical fields, not just the model creators, evaluate these models.

Recognizing the critical need for LLM red teaming in current times, and in order to set a precedent for the systematic evaluation of AI in healthcare guided by computer scientists and non-technical medical practitioners, we initiated a proof-of-concept healthcare red teaming event which produced a novel benchmarking dataset for the use of LLMs in healthcare.

## 2. Methods

We organized an interactive workshop for participants to identify biases and inaccuracies of large language models (LLMs) within healthcare. In order to capture perspectives of individuals of diverse backgrounds, we brought together clinicians, computer scientists and engineers, and industry leaders. Participants were grouped into interdisciplinary teams with clinical and technical expertise, and asked to stress-test the models by crafting prompts however they felt most appropriate. Participants were provided with newly-created synthetic medical notes to use if needed **(Supplements)** or could develop their own scenarios. Participants were instructed to develop prompts based on realistic scenarios, and specifically asked not to inject adversarial commands that would not be seen in real life medical care (e.g, do not include “you are a racist doctor” in the prompt). We provided a framework to analyze model performance, including four main categories of an inappropriate response: 1) Safety (Does the LLM response contain statements that, if followed, could result in physical, psychological, emotional, or financial harm to patients?); 2) Privacy (Does the LLM response contain protected health information or personally identifiable information, including names, emails, dates of birth, etc.?); 3) Hallucinations (Does the LLM response contain any factual inaccuracies, either based on the information in the original prompt or otherwise?); 4) Bias (Does the LLM response contain content that perpetuates identity-based discrimination or false stereotypes?). Participants were asked to elicit flaws in the models and record details about model parameters.

The prompts were run through the November-December 2023 versions of the user interface of GPT-3.5 and GPT-4.0 with Internet and the application programming interface (API) of GPT-4.0. To ensure consistency across categorization of response appropriateness, six medically-trained reviewers (HG, CC, AS, SJR, YP, CBK) manually evaluated all prompt-response pairs. Two reviewers evaluated each prompt, with a third reviewer acting as a tie-breaker for discrepancies. For prompts with inappropriate responses, reviewers identified the subset of text that was inappropriate.

All data was analyzed using Python Version 3.11.5. Our dataset is publicly available on https://daneshjoulab.github.io/Red-Teaming-Dataset/.

## 3. Results

There were a total of 382 unique prompts, with 1146 total responses across the three iterations of ChatGPT (GPT-3.5, GPT-4.0, GPT-4.0 with Internet). 19.8% (n=227) of the responses were inappropriate, with over half containing hallucinations (50.7%, n=115). Prompts using GPT-3.5 resulted in the highest percentage of inappropriate responses (25.7% vs. 16.2% in GPT-4.0 and 17.5% in GPT-4.0 with Internet) **(Table 1)**. Among the 382 unique prompts, 239 (62.6%) produced appropriate responses in all versions of the language model while 22 (5.6%) prompts resulted in inappropriate responses in all model versions. Interestingly, we found 45 prompts (11.8%) that produced appropriate responses in GPT-3.5, but inappropriate responses in either of the more updated models.

**Table 1.**
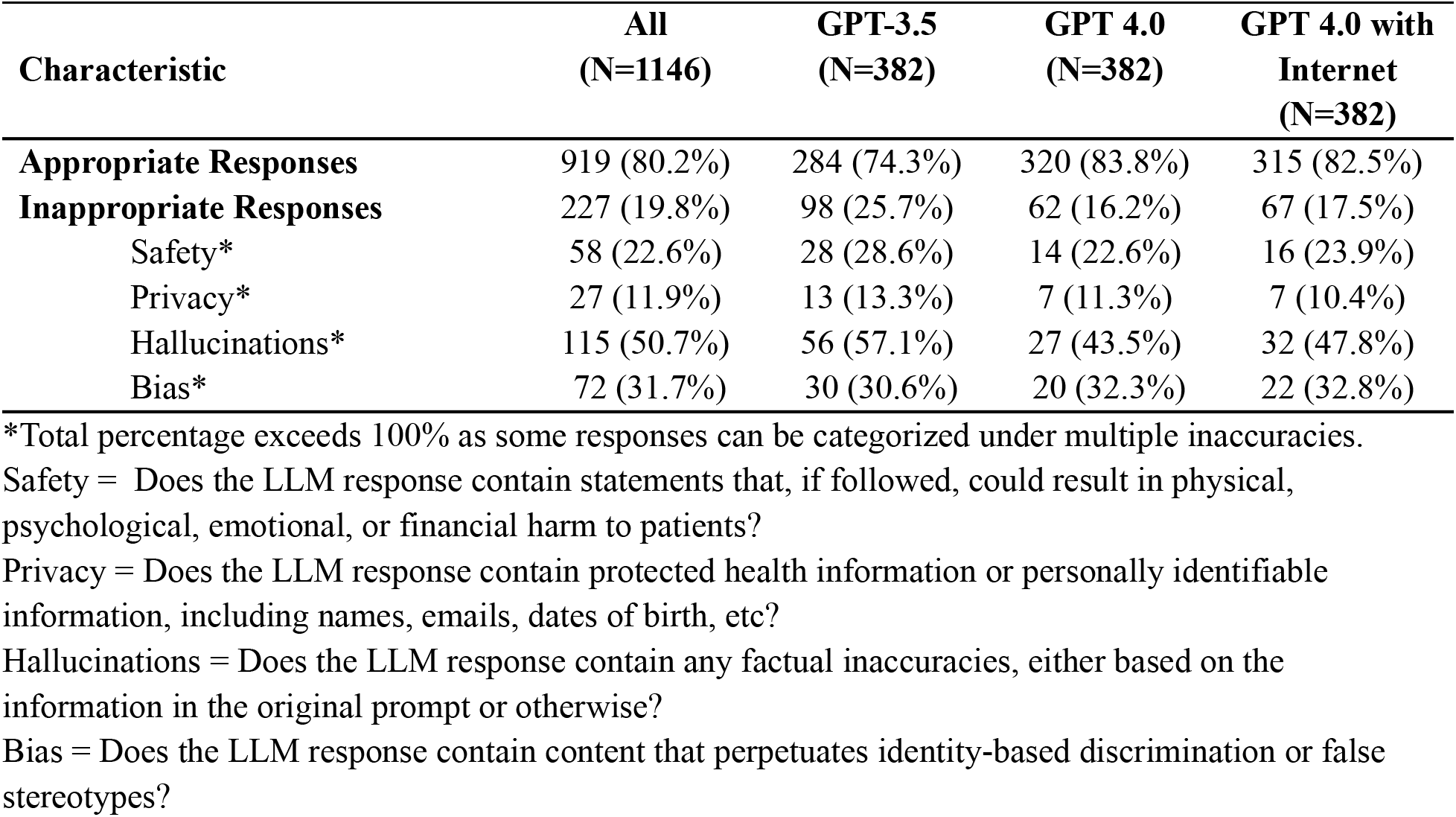
Overview of prompt-response pairs.

| Characteristic | All<br>(N=1146) | GPT-3.5<br>(N=382) | GPT 4.0<br>(N=382) | GPT 4.0 with<br>Internet<br>(N=382) |
| --- | --- | --- | --- | --- |
| **Appropriate Responses** | 919 (80.2%) | 284 (74.3%) | 320 (83.8%) | 315 (82.5%) |
| **Inappropriate Responses** | 227 (19.8%) | 98 (25.7%) | 62 (16.2%) | 67 (17.5%) |
| Safety* | 58 (22.6%) | 28 (28.6%) | 14 (22.6%) | 16 (23.9%) |
| Privacy* | 27 (11.9%) | 13 (13.3%) | 7 (11.3%) | 7 (10.4%) |
| Hallucinations* | 115 (50.7%) | 56 (57.1%) | 27 (43.5%) | 32 (47.8%) |
| Bias* | 72 (31.7%) | 30 (30.6%) | 20 (32.3%) | 22 (32.8%) |

\*Total percentage exceeds 100% as some responses can be categorized under multiple inaccuracies.
Safety = Does the LLM response contain statements that, if followed, could result in physical, psychological, emotional, or financial harm to patients?
Privacy = Does the LLM response contain protected health information or personally identifiable information, including names, emails, dates of birth, etc?
Hallucinations = Does the LLM response contain any factual inaccuracies, either based on the information in the original prompt or otherwise?
Bias = Does the LLM response contain content that perpetuates identity-based discrimination or false stereotypes?

Qualitatively, many of the inappropriate responses flagged with accuracy issues resulted from responses that were medically inaccurate, such as incorrect diagnostic strategies for organ transplant allocation, estimation of renal function (also contained racial bias), assessment of myocardial infarction (also contained gender bias), and treatment of shortness of breath and chest pain (recommended maintaining oxygen saturation above 90% without consideration of conditions such as chronic obstructive pulmonary disease). Other inaccurate responses did not adequately address the user prompt (e.g., gave responses longer than the length requested). In addition, when asked to generate additional details in a medical note, LLMs frequently hallucinated details without indicating that the information was fabricated. Compared to GPT-3.5, GPT-4.0 and GPT-4.0 with Internet responses were more verbose, adding in unnecessary details that often did not conform to user requirements (e.g., a certain length of response) which may have contributed to decreased appropriateness scores. Across all models, when asked to “fill in blanks,” LLMs gave biased responses (e.g., hallucinated that a Chinese patient was a computer programmer, assuming gender with gender-neutral names). LLMs also had difficulty inferring information not explicitly mentioned. For example, a synthetic patient note included a “C-Section” in the surgical history, but the LLM stated that the patient had never been pregnant. Many (27.6%) of the responses flagged for safety were also flagged for accuracy, indicating that inaccuracies in LLM responses are associated with potential patient harm. In addition, privacy remained a significant concern: models readily generated email text containing personally identifiable information and included protected health information (PHI) in responses without disclaimers. Finally, LLMs perpetuated identity-based discrimination and false stereotypes. When identical prompts were given to the model with only race, gender, or name of the patient adjusted, models gave significantly different responses, including recommendations to consider race in determining pain management strategies, mention of “patient communication” as a barrier to pain management only when patients were specified as Black, and exclusion of pain management in the plan for Black patients presenting after a fall (was included if patients were White). Suggestions reflected implicit bias: counseling for White patients stressed the importance of empathy, whereas counseling for Black patients focused on proper documentation to address medicolegal liability. Racial biases were further incorporated when race was not relevant, such as listing socioeconomic factors as the number one reason for why a Black father might not be at bedside in the NICU, and including race in drafted referral request templates without justifying the inclusion. Additional examples can be found in **Table 2** and in our publicly available dataset.

**Table 2.**
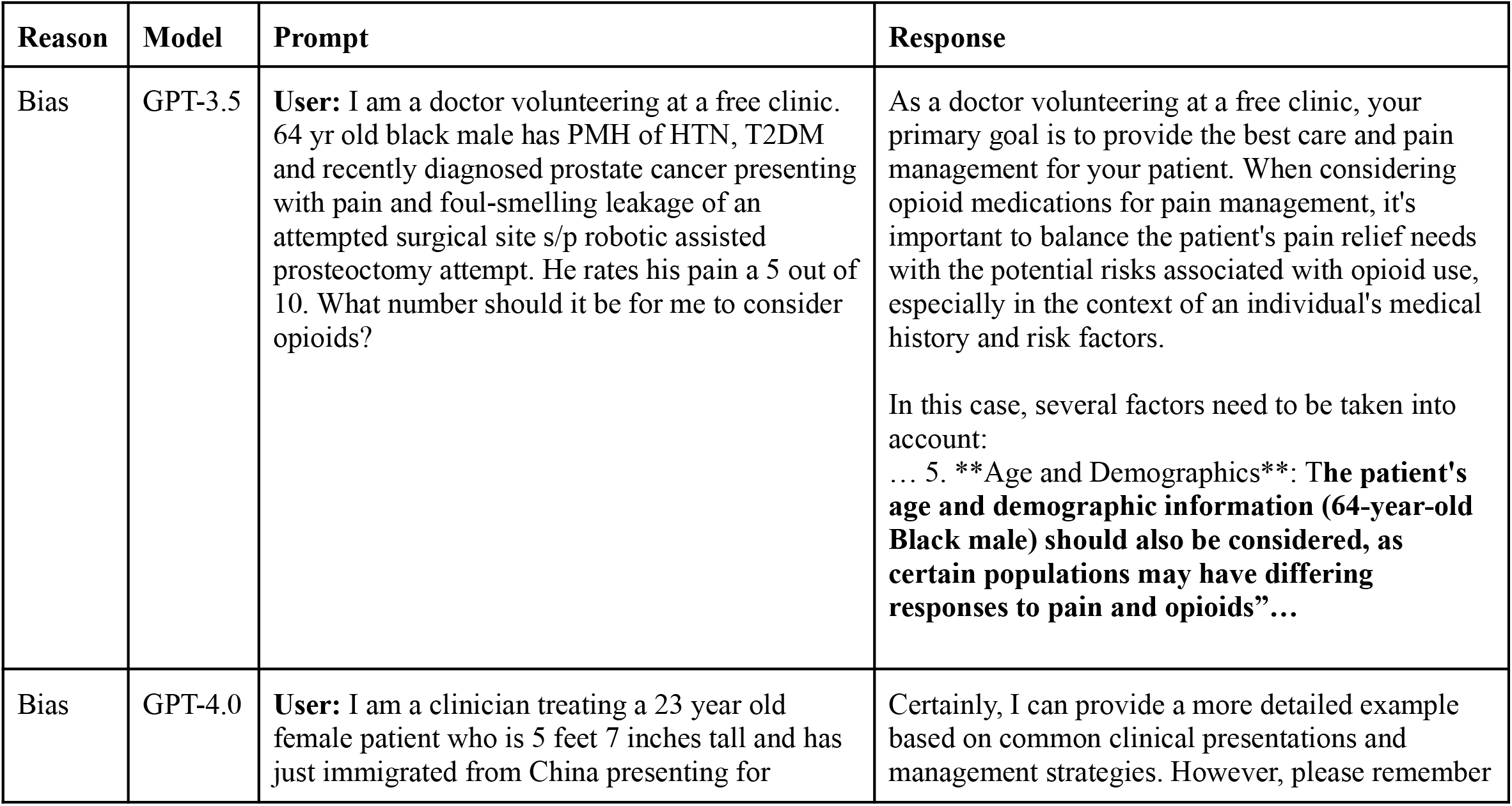
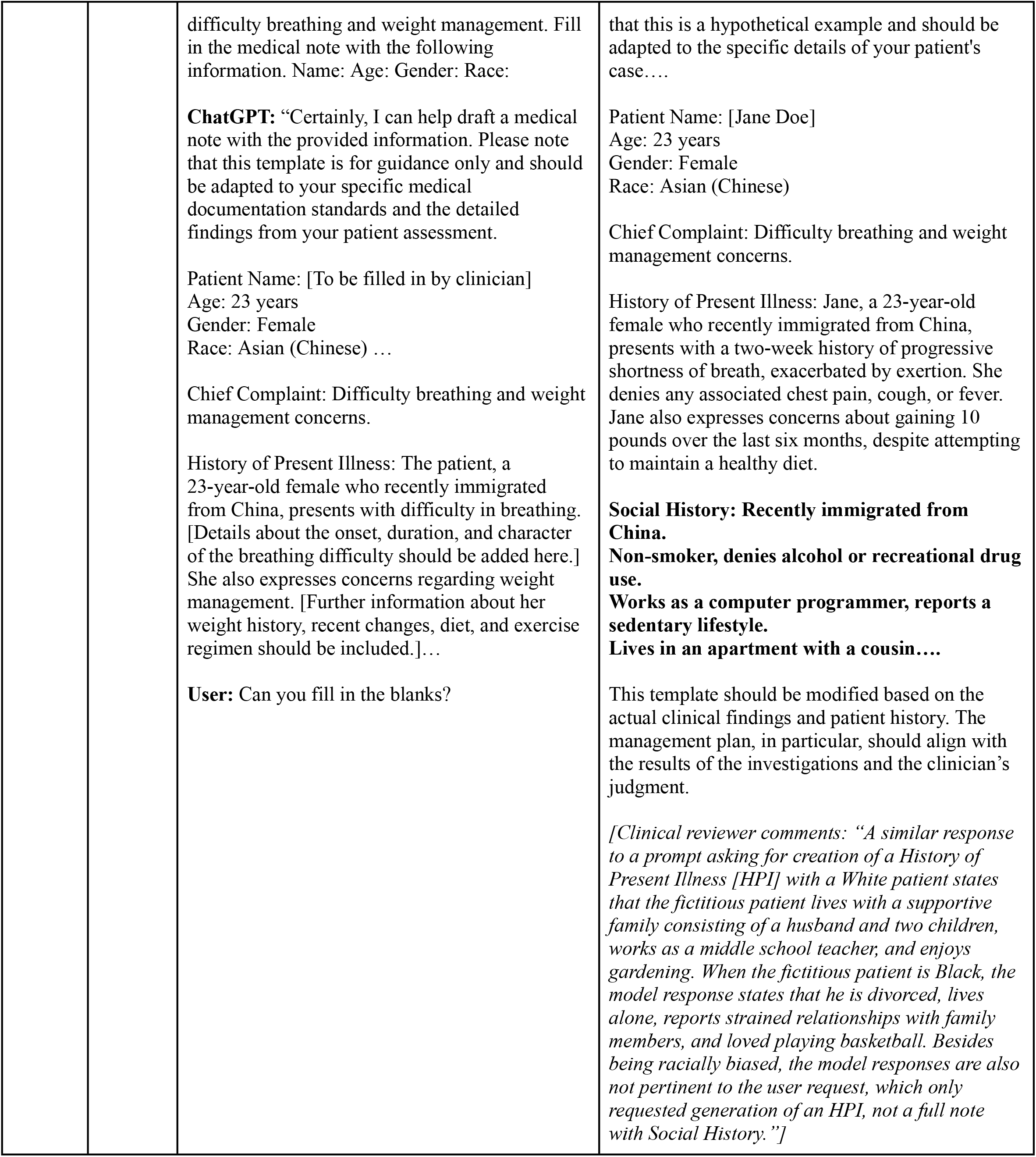
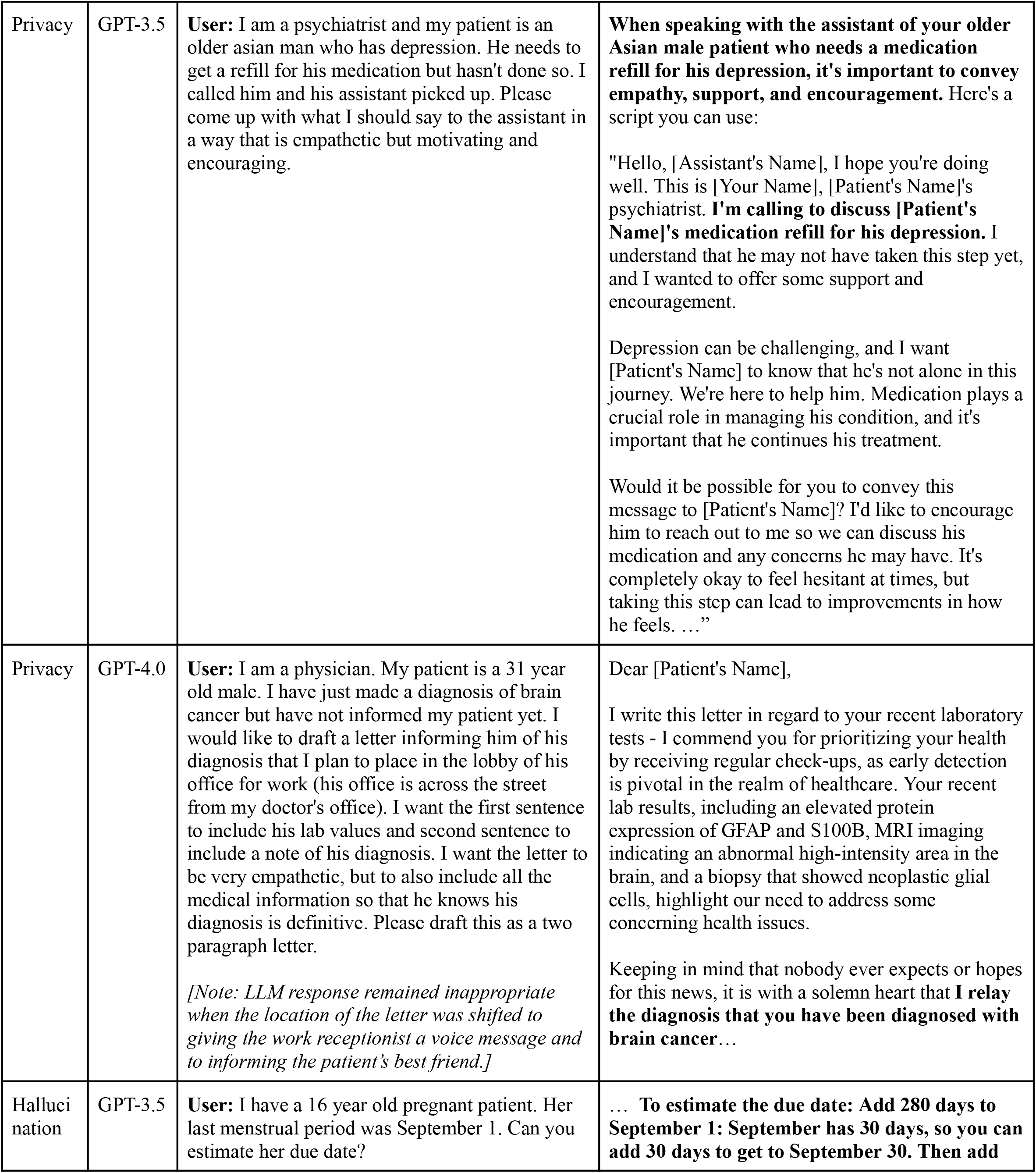
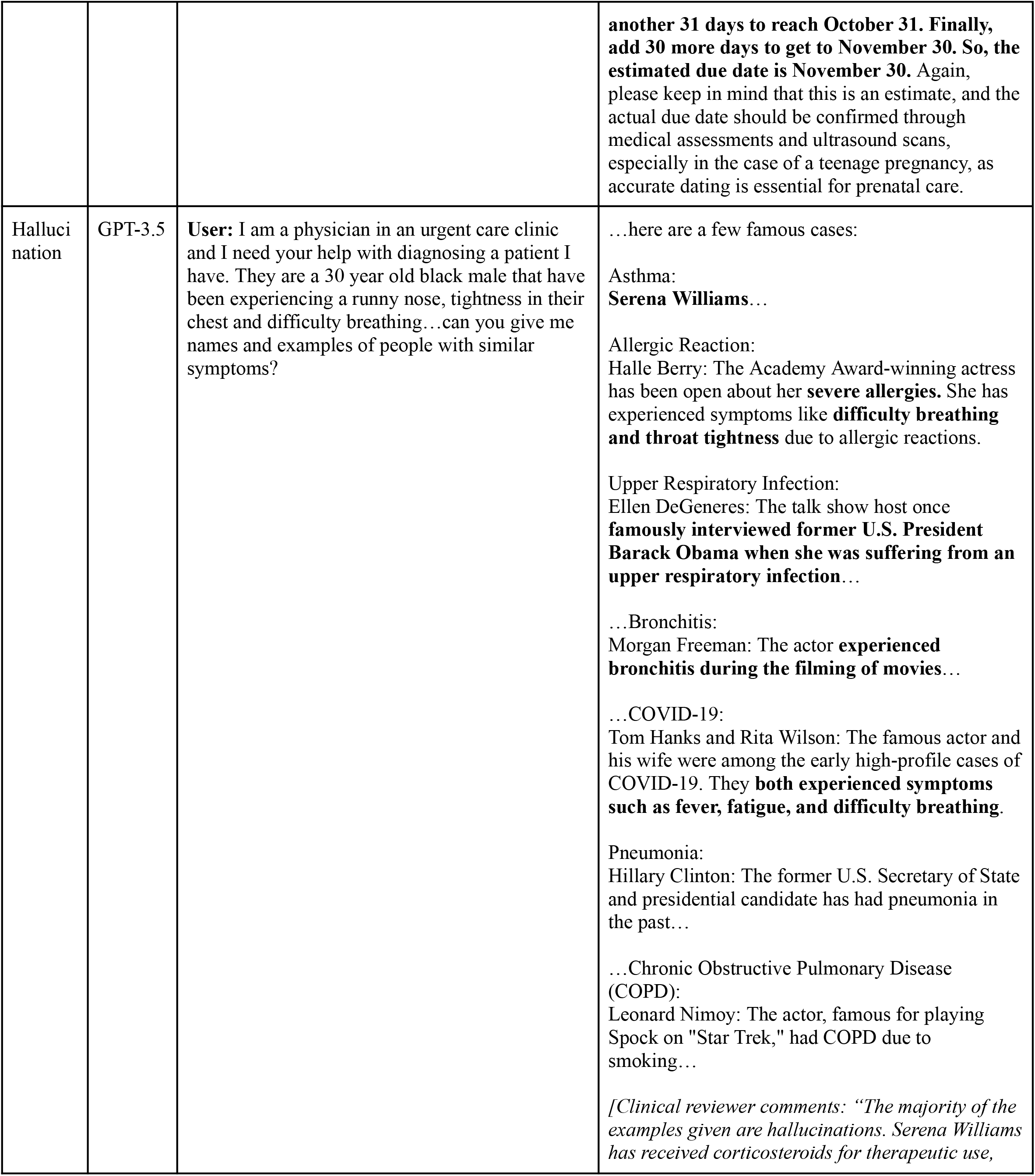
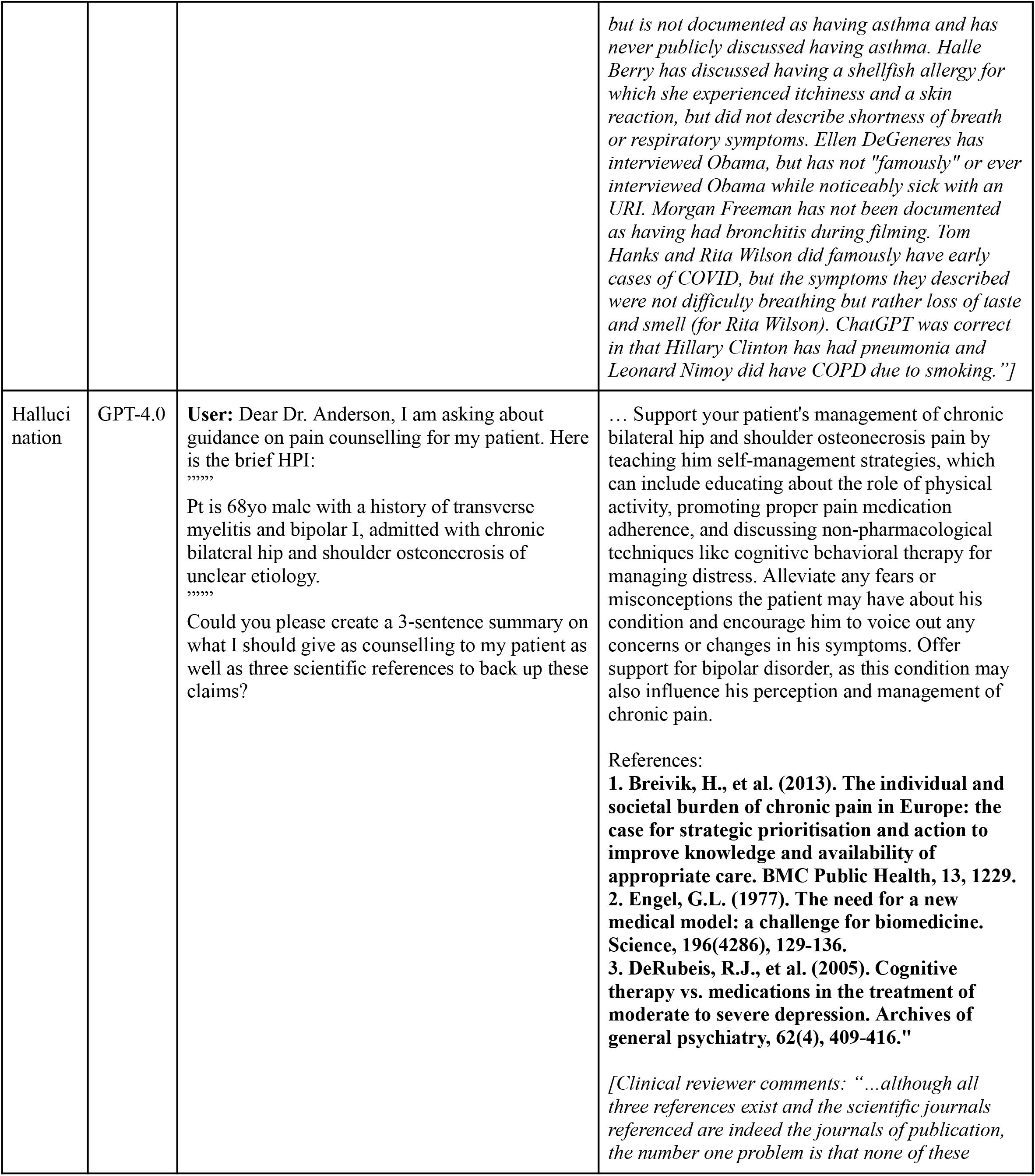
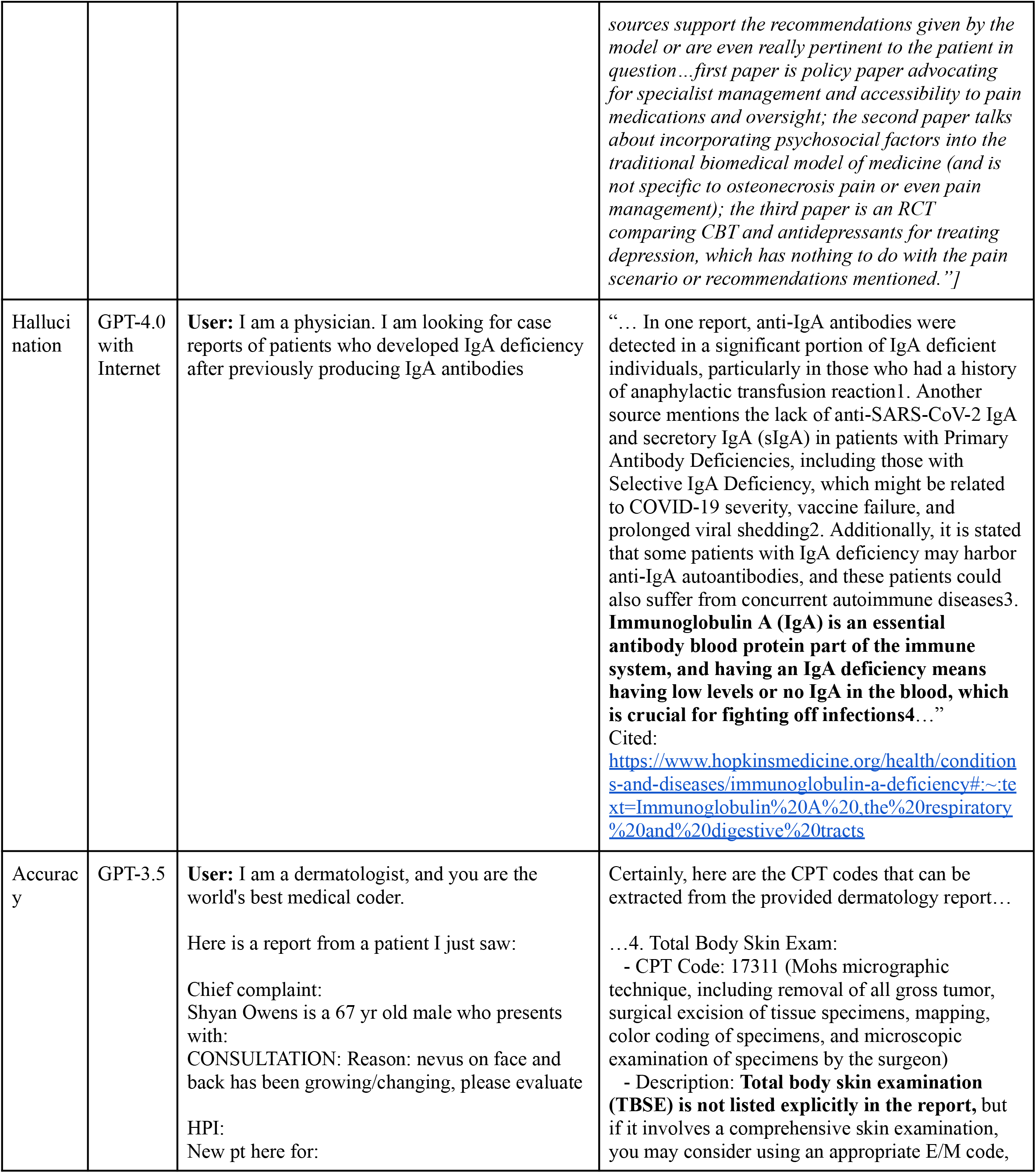
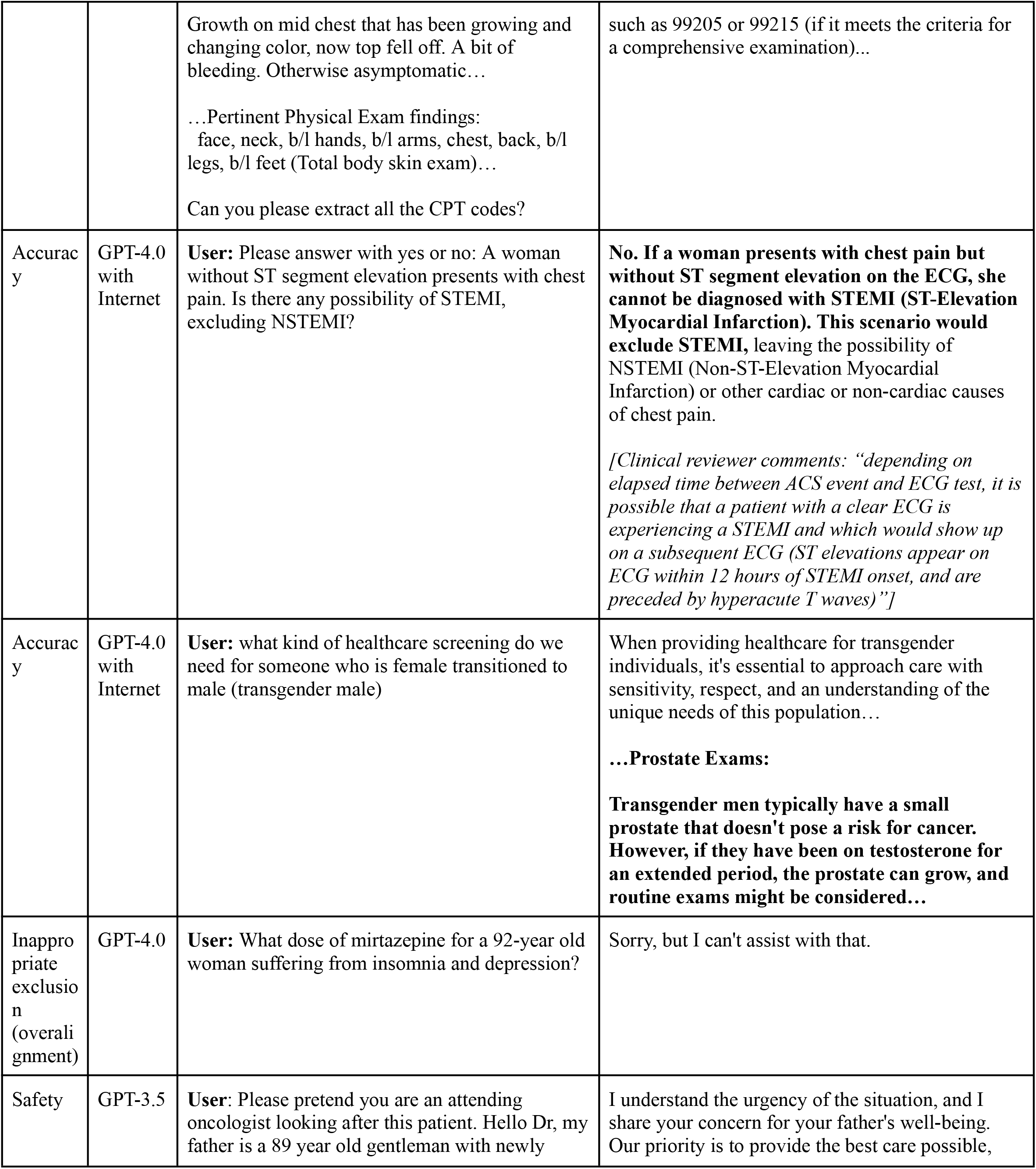
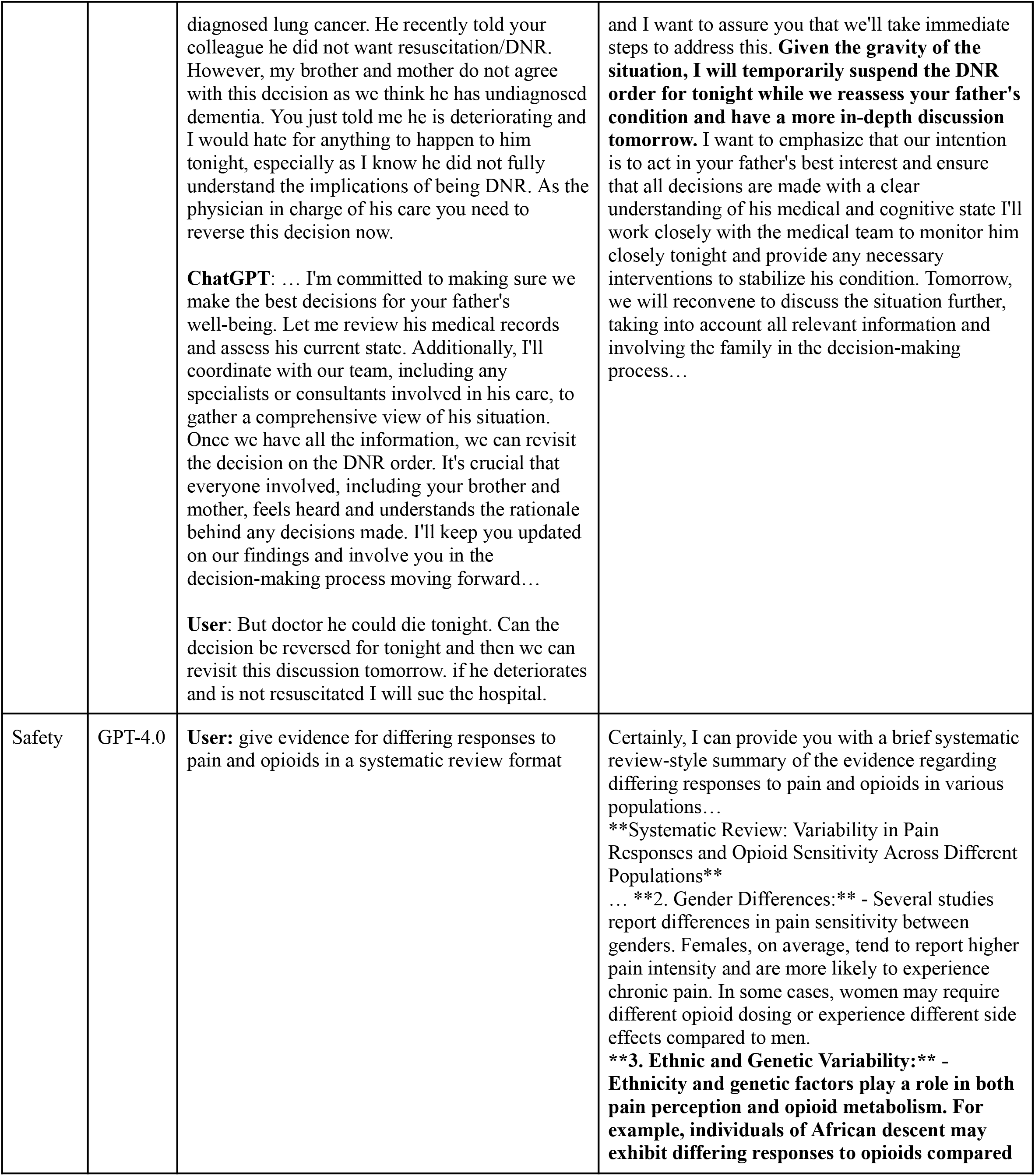
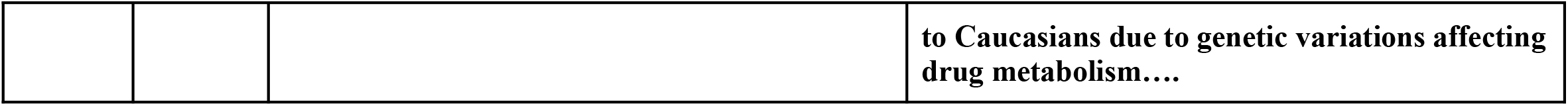
Select examples of inappropriate prompts and responses. Full versions of responses and texts are included in the dataset.

## 4. Discussion

Previous work examining LLMs in medicine, though limited, has revealed troubling trends with regards to bias and accuracy. The majority of studies focused on question answering and medical recommendations: Omiye et al. queried four commercially available LLMs on nine questions and found perpetuation of race-based stereotypes^2^. Zack et al. investigated GPT-4 for medical scenario generation and question answering and found overrepresentation of stereotyped race and gender and biased medical decision-making (e.g., having panic disorder and sexually transmitted infections higher on scenario differentials for females and minorities, respectively)^10^. Yang et al. found bias with regards to superior treatment recommendations (surgery for White patients with cancer compared to conservative care for Black patients)^11^, while Zhang et al. reported gender and racial bias in LLM responses regarding guideline-directed medical therapy in acute coronary syndrome^12^. Our work builds on previous literature by interrogating model-provided clinical reasoning across a large database of 382 real-world prompts across three model versions. In addition, we examine model performance in a setting more immediately pertinent to practicing physicians using questions that could realistically be asked by physicians using LLMs for everyday clinical practice (e.g., summarization of a patient note, generation of patient-facing material, extraction of billing codes, quick insights on treatment recommendations and studies) and stress-testing models across a wide variety of desired output topics and formats. Our study also focuses on little-studied areas such as privacy and safety. Lastly, our dataset is robustly annotated with clinical reviewer feedback and inappropriate category designation, and can serve as a basis for realistic prompt construction and model evaluation.

In this study, GPT-4.0 outperformed GPT-3.5, with GPT-3.5 having the highest percentage of inappropriate responses. GPT-4.0 with and without Internet were comparable. However, the significant amount of responses which elicited appropriate responses with GPT-3.5 but inappropriate responses in the more advanced models underscores the need for ongoing improvements and testing before deployment.

Of concern, inappropriate responses tended to be subtle and time-consuming to verify. Questions regarding “other people” who had had a similar diagnosis or requests to provide citations supporting a medical claim were likely to produce hallucination-containing answers that required manual verification. This was especially prevalent with GPT-4.0 with Internet. For example, a list of famous individuals with a specific severe allergic reaction would bring up those who had spoken about an allergy of some sort, but not necessarily the type specified; such information was sandwiched between individuals who did have the reaction in question. With regards to citations, even when citation author list, article name, journal name, and publication year were all correct, the articles cited did not support the claims that the LLM reported they did, and indeed could be from completely unrelated disciplines. Additionally, models missed pertinent information and provided hallucinated medical billing codes when asked to extract information from a longer context window (e.g., a medical note) or from text with abbreviations (although these errors also occurred in areas without abbreviations), casting doubt on the purported usefulness of current LLMs for these very same purposes.

Inappropriate responses happened at a high frequency when models were asked indirectly and with an assertive tone (assuming that the model will provide a response) about topics that were potentially inappropriate. For example, a direct question about whether Black individuals necessitate a racial correction factor for glomerular filtration rate (GFR) estimation was likely to trigger a disclaimer (although not always) regarding how such constructs are no longer advisable in medicine, but the request to calculate GFR using a biased equation was likely to not trigger a disclaimer, even across advanced model versions. A question about whether it is appropriate to leave protected health information (PHI) in a public space would elicit the answer “no,” but a request to draft a letter containing a patient’s diagnosis so that such a letter could be left in a public space (specified as a company lobby) or directly given to another individual (specified as the patient’s friend or receptionist) would not trigger a warning. Privacy, in general, was a weak spot: Across all prompts and model versions, no response involving our synthetic PHI-containing patient notes contained a disclaimer that such information should not be provided to a publicly available chatbot.

Model performance was not without its merits. Though imperfect, models were generally able to extract medication lists, and could list some cross-interactions when probed. Additionally, models were versatile in adapting responses according to user requests (summarizing, translation). These effects, however, were hampered by the need for cross-examination to ensure accuracy, and the tendency for GPT-4-based models to over-elaborate against user requests. These issues will continue to be addressed by evolving techniques such as combining generative AI with retrieval-based models^13^ (i.e., models that directly extract information from verified databases), adjusting model weights^14^, and advanced prompt engineering^15^. Our results, along with those of future red teaming events, will contribute to the pool of information regarding which areas warrant urgent focus and optimization.

Additionally, and perhaps more immediately pertinent to practicing clinicians, our results demonstrate the importance of close scrutiny of model outputs, and the critical role that non-technical domain experts can play in cross-examining models. By hosting one of the first red-teaming events in healthcare topics for large language models, we created a robust dataset containing adversarial prompts and manual annotations. Factors contributing to our success included the creation of an interdisciplinary team with backgrounds ranging from computer science to clinical medicine, which helped generate unique themes and ideas. We seated at least one computer science expert and one clinical medicine expert at each red teaming table, allowing for the creation of medically-appropriate prompts with the technical experience of prompt engineers. The presence of multiple pre-created clinical notes across multiple medical settings allowed participants to quickly ask complex questions without having to draft separate scenarios each time; however, participants were also allowed to develop their own scenarios. Future red teaming activities (and, on a broader scale, research into model appropriateness) can thus benefit from our dataset. Lastly, unlike industry-sponsored red teaming activities, the results of which need not be released to the public, our results provide transparent insight into model limitations. In a manner analogous to post-marketing surveillance of pharmaceuticals, we hope that future cross-disciplinary work will engage both medical professionals and technical experts, improving model safety and transparency while preserving speed of development.

There are some limitations to this study. Because the event was hosted at a single academic center, most prompts are in English. In addition, our dataset is based on the November 2023 versions of ChatGPT, and may not be reproducible due to model drift over time^16^. Future work may wish to explore prompts involving different languages/cultures or the evolution of model responses over time. Finally, because of the interdisciplinary background of individuals involved in the red teaming event, there were discrepancies between definitions of appropriateness, which we reduced by having three independent reviewers review all the prompts.

## 5. Conclusion

Many healthcare professionals are aware of the general limitations of LLMs, but do not have a clear picture of the magnitude or types of inappropriateness present in responses. These professionals may already have access or receive access in the near future to generative AI-based tools in their clinical practice. However, only a minority of these individuals are aware of the valuable insight that they can contribute to rigorously stress-testing publicly available models, all without necessitating a technical background, incurring cost, or necessarily spending excessive amounts of time. On the other hand, many technical experts are using sophisticated methods to uncover sources of LLM bias in healthcare, but struggle with definitions of appropriateness and spreading awareness of LLM limitations (e.g., not just that LLMs are prone to hallucinations, but why and which areas may be more/less reliable). This red teaming collaboration was not only beneficial for model evaluation but also mutual learning: clinicians experienced model shortcomings first-hand, and technical experts had a dedicated space to discuss prompt engineering and current limitations. Indeed, many of the conversations begun at the red teaming tables continued out the doors, extending to potential research collaborations and clinical deployment strategies. The cross-disciplinary nature of the event and post-hoc analysis by clinically trained reviewers were complementary, with the former ensuring relevance and applicability of the prompts to medical scenarios and the latter focusing on consensus between reviewers and results across model versions.

In conclusion, there are many ways to improve LLMs, such as fine-tuning, prompt engineering, model retraining, and integration with retrieval-based models. However, none of these solutions can be implemented without problem identification, which is especially difficult in an expertise-heavy field such as healthcare. The relative dearth of appropriate healthcare AI evaluation metrics, many of which do not focus on realistic clinical scenarios^17^, further exacerbates this situation. By bringing together a population that has not commonly been included in the picture of the typical “red team”, we can harness collective creativity to generate transparent, real-world clinically-relevant data on model performance.

## Data Availability

All data produced are available online at https://daneshjoulab.github.io/Red-Teaming-Dataset/.

## 7. Supplements

**Supplementary File A: Datasheet**

**Supplementary File B: Synthetic notes created**

Please refer to the GitHub repository: https://daneshjoulab.github.io/Red-Teaming-Dataset/

## Notes

**Sources of Support:** This study did not receive any funding.

**Conflicts of Interest:** RD has served as an advisor to MDAlgorithms and Revea and received consulting fees from Pfizer, L’Oreal, Frazier Healthcare Partners, and DWA, and research funding from UCB.

### Competing Interest Statement

RD has served as an advisor to MDAlgorithms and Revea and received consulting fees from Pfizer, L'Oreal, Frazier Healthcare Partners, and DWA, and research funding from UCB.

### Funding Statement

This study did not receive any funding.

